# Neurodevelopmental and psychiatric conditions in 600 Swedish Children with the ARFID phenotype

**DOI:** 10.1101/2024.05.16.24307471

**Authors:** Manda Nyholmer, Marie-Louis Wronski, Liv Hog, Ralf Kuja-Halkola, Paul Lichtenstein, Sebastian Lundström, Henrik Larsson, Mark J. Taylor, Cynthia M. Bulik, Lisa Dinkler

## Abstract

**Objective:** Avoidant restrictive food intake disorder (ARFID) is a feeding and eating disorder characterized by extremely restricted dietary variety and/or quantity resulting in serious consequences for physical health and psychosocial functioning. ARFID often co-occurs with neurodevelopmental conditions (NDCs) and psychiatric conditions, but previous data are mostly limited to small clinical samples examining a narrow range of conditions. Here, we examined NDCs and psychiatric conditions in a large, population-based group of children with ARFID.

**Method:** In a sample of 30,795 children born 1992–2008 in Sweden, ARFID was assessed using parent reports and clinical diagnoses from national health registers. Parents further reported symptoms of NDCs and psychiatric conditions at child age 9 or 12 years. We conducted regressions for symptom scores and screening diagnoses (identified using validated cut offs) on ARFID and examined interactions with sex.

**Results:** Children with ARFID had significantly increased odds of all 17 screening diagnoses with odds ratios ranging from 3.3 for visual hallucinations to 13.7 for autism (all p<.0001). The most common NDCs were oppositional defiant disorder (19.4%), ADHD (16.9%), tic disorders (14.8%), and autism (12.1%). Among psychiatric conditions, separation anxiety disorder (29.0%) and sleep problems (20.0%) had the highest prevalence. We did not find any sex-specific differences in co-occurring conditions.

**Conclusion:** This study highlights the co-occurrence of a broad range of NDCs and psychiatric conditions with ARFID in a large, non-clinical cohort. Our findings underscore that children with ARFID face significant burden from multiple co-existing conditions which should be considered during assessment and treatment.

## Introduction

Eating disorders are serious, potentially life-threatening psychiatric conditions associated with clinically significant impairment, high disability, and increased mortality.^1^ Co-occurring neurodevelopmental conditions (NDCs) and psychiatric conditions seem to be present in the majority of individuals diagnosed with an eating disorder.^2^ However, less is known about the comorbidity of NDCs and psychiatric conditions with the relatively recently introduced eating disorder diagnosis avoidant/restrictive food intake disorder (ARFID).

ARFID was formally recognized as a psychiatric diagnosis in DSM-5,^3^ and subsequently included in ICD-11.^4^ The disorder is characterized by an extremely restricted *variety* and/or *quantity* of food intake that is unrelated to weight concern or body image disturbance and that leads to severe and potentially life-threatening physical, nutritional, and psychosocial consequences.^3^ Occurring in 1–2% of all children and adults,^5,6^ the prevalence of ARFID is comparable to that of autism and anorexia nervosa.^7,8^ Unlike other eating disorders, the distribution of males and females with ARFID has been reported to be approximately equal.^6,9^ On average, ARFID has an earlier age of onset than other eating disorders, though it occurs at all ages and is sometimes life-long.^3,10^

ARFID commonly co-occurs with NDCs and mental health problems.^11–13^ Existing studies indicate that between 45% and 82% of youth with ARFID have a co-existing psychiatric condition.^14,15^ Most commonly reported are anxiety disorders (9%–72%), especially generalized anxiety disorder (GAD), and autism (8%–55%).^12^ Depressive symptoms are also common (7–33%), but less frequent than in other eating disorders.^12^ Some studies also reported an elevated prevalence of attention deficit hyperactivity disorder (ADHD; 3–23%),^16,17^ learning difficulties/cognitive impairment,^16–18^ and obsessive-compulsive disorder (OCD).^17,18^

Little data exist for other NDCs and psychiatric conditions. Much of the available data stem from low-powered, small samples, often without a control group, leading to broad/nonspecific ranges of estimates. Prevalence estimates of co-occurring psychiatric conditions also vary considerably depending on the specific population studied (e.g., adults vs. children, inpatient vs. outpatient, eating disorder treatment centers vs. pediatric feeding clinics). Additionally, previous data almost exclusively stem from clinical samples and therefore do not provide information on the large group of people with ARFID who do not seek clinical care.

Given the sex differences observed in the prevalence of NDCs and psychiatric conditions,^19^ it is important to investigate whether these sex differences are also present in individuals with ARFID. Whether patterns of comorbidity in ARFID differ by sex is currently unclear, because previous studies examining this issue have not been sufficiently powered^11^ or focused on a very specific subgroup of ARFID.^20^ One study found that girls with a diagnosed feeding disorder before age 3 were at greater risk of developing later emotional/behavioral conditions and intellectual disability, but not of having ADHD and autism.^20^

The frequent comorbidity in ARFID engenders complex and heterogeneous clinical presentations, complicating differential diagnosis and treatment planning.^21^ A deeper understanding of the patterns of co-occurring NDCs and psychiatric conditions in ARFID, and potential sex differences in such patterns, could provide insights into ARFID etiology and potential shared underlying mechanisms, which, in turn could facilitate the refinement of diagnostic schema and tailoring of treatment.

In the current study, we examined a broad range of NDCs and psychiatric conditions as both continuous and dichotomous outcomes in a large group of children with the ARFID phenotype from the general child population. In line with the existing literature, we hypothesized that children with ARFID would have higher symptom scores and higher odds of screening diagnosis compared with children without ARFID on most of the studied domains, especially, autism, ADHD, and anxiety. We did not have specific hypotheses regarding sex differences.

## Method

### Participants

The current study included 30,795 children (49.2% female) born 1992–2008 from the Child and Adolescent Twin Study in Sweden (CATSS). CATSS is a nationwide, longitudinal study, where families of all twins born in Sweden since July 1, 1992 are invited to participate when the twins are 9 years old (the first three cohorts were contacted at age 12 years).^22^ Here we used parent reports of their children’s neurodevelopment and mental health from the first CATSS wave (at child age 9/12 years; response rate ∼69%). CATSS has ethical approval from the Regional Ethics Review Board in Stockholm, Sweden (Dnrs 03-672, 2010/597-31/1, 2010/322-31/2). Informed assent was obtained from parents.

### Assessment of ARFID

We previously constructed a composite measure to identify the ARFID phenotype in this sample by drawing on all available information relevant to the DSM-5 ARFID criteria.^5^ This included parent reports, diagnostic and procedural codes from the National Patient Register, and prescribed drugs from the National Prescribed Drug Register. To be identified as having ARFID, children needed to meet DSM-5 ARFID criterion A (avoidant/restrictive eating leading to clinical consequences, e.g., low weight, nutritional deficiencies, and psychosocial impairment) and DSM-5 ARFID criterion C (eating disturbance not attributable to anorexia nervosa, bulimia nervosa, or body image disturbance) between the age of 6 and 12 years. Data to assess DSM-5 ARFID Criterion B (eating disturbance not explained by lack of available food or associated culturally sanctioned practices) were not available. In line with our previous studies on the same sample,^5,11^ we did *not* exclude individuals based on DSM-5 ARFID criterion D, as it is difficult to ascertain in an epidemiological context whether the eating disturbance could be explained by another somatic or mental condition. A detailed overview of the extracted information used to identify each ARFID criterion can be found elsewhere.^5,11^

### Assessment of outcomes

NDCs and psychiatric conditions were measured using the *Autism-Tics, ADHD, and other Comorbidities* (A-TAC) inventory,^22,23^ the *Screen for Child Anxiety Related Emotional Disorders* (SCARED),^24^ and the *Short Mood and Feelings Questionnaire* (SMFQ).^25^ All outcomes were parent-reported and were applied both continuously and dichotomously by using cutoff values to create screening diagnoses. **Table 1** shows a detailed overview of the applied scales, scoring, cutoff values, and psychometric data.

**Table 1.** Scales assessing psychiatric and neurodevelopmental problems (outcomes)

| Scale | No. of items | Theoretical range | Included as continuous outcome | CA in the current sample | Included as categorical outcome | Cutoff ≥ | Sensitivity/Specificity<sup>a</sup> | PPV /NPV<sup>a</sup> |
| --- | --- | --- | --- | --- | --- | --- | --- | --- |
| **Neurodevelopment (A-TAC)** |  |  |  |  |  |  |  |  |
| ADHD | 19 | 0-19 | X | .92 | X | 12.5 | .28 / .99 | .38 / .98 |
| Inattention | 9 | 0-9 | X | .90 |  | <sup>b</sup> |  |  |
| Impulsivity/hyperactivity | 10 | 0-10 | X | .87 |  | <sup>b</sup> |  |  |
| Autism | 17 | 0-17 | X | .84 | X | 8.5 | .30 / .99 | .33 / .99 |
| Social communication | 12 | 0-12 | X | .78 |  | <sup>b</sup> |  |  |
| RRBI | 5 | 0-5 | X | .72 |  | <sup>b</sup> |  |  |
| Oppositional defiant disorder | 5 | 0-5 | X | .75 | X | 3 | .57 / .97 | .02 / 1.00 |
| Conduct disorder | 5 | 0-5 | X | .62 | X | 2 | .31 / .99 | .09 / 1.00 |
| Perception problems | 4 | 0-4 | X | .58 | X | 1.5<sup>b</sup> | ---- | ---- |
| Learning disorders | 3 | 0-3 | X | .74 | X | 3 | .39 / .93 | .22 / .99 |
| Tic disorders | 3 | 0-3 | X | .61 | X | 1.5 | .52 / .97 | .05 / 1.00 |
| Sleep problems | 3 | 0-3 | X | .55 | X | 1<sup>b</sup> | ---- | ---- |
| Obsessive compulsive disorder | 2 | 0-2 |  | .65 | X | 1 | .30 / .98 | .04 / 1.00 |
| Developmental coordination disorder | 1 | 0-1 |  | ---- | X | 1 | .32 / .98 | .06 / 1.00 |
| Visual hallucinations | 1 | 0-1 |  | ---- | X | 0.5<sup>b</sup> | ---- | ---- |
| **Depression (SMFQ)** |  |  |  |  |  |  |  |  |
| Total score | 13 | 0-26 | X | .83 | X | 8 (girls)<br>7 (boys) | .67/.83<br>.63/.65 | ---- |
| **Anxiety (SCARED)** |  |  |  |  |  |  |  |  |
| Total score (any anxiety disorder) | 41 | 0-82 | X | .88 | X | 14 | .68/.61 | ---- |
| Panic disorder | 13 | 0-26 | X | .60 | X | 4 | .83/.83 | ---- |
| Generalized anxiety disorder | 9 | 0-18 | X | .81 | X | 8 | .56/.80 | ---- |
| Separation anxiety disorder | 8 | 0-16 | X | .73 | X | 5 | .79/.80 | ---- |
| Social phobia | 7 | 0-14 | X | .86 | X | 9 | .75/.88 | ---- |
| School avoidance | 4 | 0-8 | X | .63 |  | <sup>b</sup> |  |  |

A-TAC was available for birth years 1992–2010; scoring: No (0), Yes, to a certain degree (0.5), Yes (1). SCARED and SMFQ were available for birth years 1998–2010; scoring: No (0), Yes, to a certain degree (1), Yes (2). Scales including <3 items were not included as continuous outcomes. <sup>a</sup>References: Mårland et al. 2017 (A-TAC); Jarbin et al. 2020 (SMFQ), Ivarsson et al. 2018 (SCARED). <sup>b</sup>No established cut-off values available as there is no corresponding clinical diagnosis. For perception problems, sleep problems, and visual hallucinations, cut-offs were chosen based on reasonable prevalence in the full sample. ADHD, attention deficit hyperactivity disorder; ARFID, avoidant restrictive food intake disorder; A-TAC, Autism-Tics, ADHD, and other Comorbidities inventory; RRBI, restricted/repetitive behavior & interests; SCARED, Screen for Child Anxiety Related Emotional Disorders; SMFQ, Short Mood and Feelings Questionnaire.

A-TAC was designed for large-scale epidemiological studies. It consists of 96 items, is based on symptom criteria and common clinical features, and is divided into modules corresponding to diagnostic domains. Several cross-sectional and longitudinal validation studies indicate good to excellent validity for many of the domains, especially autism, ADHD, oppositional defiant disorder (ODD), and learning disorders.^23,26^ For subscales without previously established cutoff values, we determined cutoff values based on a reasonable prevalence of the respective domain in the general population (i.e., full sample) for exploratory purposes.

SCARED measures anxiety symptoms over the last 3 months on five anxiety dimensions.^24^ SMFQ assesses depressive symptoms over the last 2 weeks.^25^ Both measures have been tested among Swedish psychiatric outpatients aged 6–17 years. Cutoff values identified for the parent-reported versions show acceptable sensitivity and specificity for clinical diagnoses of GAD, panic disorder, social phobia, and separation anxiety (SCARED), and for depression (SMFQ).^27,28^ In the current sample, internal consistencies (Cronbach’s α) ranged from .55 to .92. Scales including <3 items were not included as continuous outcomes (**Table 1**). A-TAC was available for the full sample. SCARED and SMFQ were only available for twins born 1998 and after (∼59.4 %, n∼18,300; 49.8% female).

### Statistical analyses

Statistical analyses were performed using R version 4.2.2.^29^ Total and subscale sum scores were calculated if responses were available for 80% of the items on each respective scale using pro-rating (i.e., the mean of the completed subscale items was imputed for the missing values on that subscale), otherwise the measurement was considered missing. To assess whether ARFID was associated with each of the outcomes, we conducted a series of linear regressions for continuous outcomes (symptom scores) and a series of logistic regressions for dichotomous outcomes (screening diagnoses, i.e., scoring above cutoff). We calculated cluster robust standard errors by treating each twin pair as a cluster to adjust for correlated observations within each pair. Sex assigned at birth (female/male) and birth year (range: 1992–2008) were included as categorical covariates in all regressions. By controlling for birth year, we simultaneously adjusted for potential differences in age at CATSS assessment (age 12 years for children born 1992–1994, age 9 years for children born 1995– 2008). To test sex effects (i.e., a difference in effect size of ARFID status on outcomes between females and males), we included an ARFID x sex interaction term in the initial regression models. Within each regression series, Benjamini-Hochberg false discovery rate (FDR)^30^ was used to adjust for multiple testing by calculating q-values (linear [logistic] regression series: adjusted across 19 [17 ] outcomes). The statistical threshold for significance was set to q < .05 for all analyses.

## Results

The current sample included 616 children with the ARFID phenotype (2.0%; 39.1% female; **Table 2**). Descriptive statistics and the results from the regressions are presented in **Table 3** (continuous outcomes/symptom scores) and **Table 4** (dichotomous outcomes/screening diagnoses). Separation anxiety disorder (29.0%) was the most prevalent condition studied among children with ARFID, followed by perception problems (22.1%), sleep problems (20.0%), and ODD (19.4%; **Table 4**). The prevalence of ADHD in the ARFID group was 16.9%, and 12.1% had autism. Of children who had available data for all screening diagnoses (n=373 with ARFID, n=17,460 without ARFID), we counted the number of screening diagnoses and determined the percentage of children with at least one screening diagnosis. Children with ARFID had on average M=2.64 screening diagnoses (SD=3.34), and 61.7% of children with ARFID had at least one screening diagnosis (comparison group: M=0.60, SD=1.41; 26.7%).

**Table 2.** Sample characteristics.

|  | **Overall**<br>N=30,795 | **ARFID**<br>N=616 | **No ARFID**<br>N=30,179 |
| --- | --- | --- | --- |
| Sex, n (%) |  |  |  |
| Male | 15,632 (50.8%) | 375 (60.9%) | 15,257 (50.6%) |
| Female | 15,163 (49.2%) | 241 (39.1%) | 14,922 (49.4%) |
| Age at CATSS assessment, n (%) |  |  |  |
| 9 years | 24,314 (79.0%) | 512 (83.1%) | 23,802 (78.9%) |
| 12 years | 6,481 (21.0%) | 104 (16.9%) | 6,377 (21.1%) |
| Birth year, n (%) |  |  |  |
| 1992-1996 | 9,469 (30.7%) | 154 (25.0%) | 9,315 (30.9%) |
| 1997-2000 | 7,739 (25.1%) | 149 (24.2%) | 7,590 (25.1%) |
| 2001-2004 | 7,327 (23.8%) | 161 (26.1%) | 7,166 (23.7%) |
| 2005-2008 | 6,260 (20.3%) | 152 (24.7%) | 6,108 (20.2%) |

ARFID, avoidant/restrictive food intake disorder; CATSS, Child and Adolescent Twin Study in Sweden

**Table 3.** Linear regression models for continuous outcomes on ARFID.

| Outcome | No ARFID |  |  | ARFID |  |  | Std. B | Unstd. B | 95% CI<sub>LL</sub> | 95% CI<sub>UL</sub> |
| --- | --- | --- | --- | --- | --- | --- | --- | --- | --- | --- |
|  | N | M | SD | N | M | SD |  |  |  |  |
| Neurodevelopmental conditions |  |  |  |  |  |  |  |  |  |  |
| Autism - Total | 30,092 | 0.79 | 1.54 | 611 | 3.28 | 3.67 | 1.48 | 2.43 | 2.13 | 2.74 |
| Autism - RRBI* | 30,103 | 0.26 | 0.61 | 613 | 1.18 | 1.39 | 1.38 | 0.90 | 0.78 | 1.01 |
| Autism - Social communication<sup>a</sup> | 30,179 | 0.54 | 1.08 | 616 | 2.11 | 2.52 | 1.33 | 1.54 | 1.33 | 1.74 |
| Perception problems | 30,179 | 0.17 | 0.46 | 616 | 0.74 | 0.98 | 1.17 | 0.56 | 0.48 | 0.64 |
| ADHD - Total | 30,068 | 2.04 | 3.07 | 608 | 5.85 | 5.39 | 1.15 | 3.67 | 3.23 | 4.11 |
| ADHD - Inattention<sup>a</sup> | 30,087 | 1.06 | 1.74 | 609 | 3.14 | 2.94 | 1.12 | 2.01 | 1.77 | 2.25 |
| Oppositional defiant disorder | 30,099 | 0.45 | 0.84 | 614 | 1.35 | 1.43 | 1.00 | 0.87 | 0.75 | 0.99 |
| ADHD - Impulsivity/hyperactivity<sup>a</sup> | 30,086 | 0.98 | 1.67 | 611 | 2.71 | 2.87 | 0.96 | 1.66 | 1.42 | 1.89 |
| Learning disorders | 30,128 | 0.28 | 0.61 | 616 | 0.80 | 1.03 | 0.82 | 0.52 | 0.43 | 0.60 |
| Conduct disorder | 30,105 | 0.10 | 0.35 | 613 | 0.36 | 0.76 | 0.69 | 0.25 | 0.19 | 0.32 |
| Tic disorders | 30,106 | 0.16 | 0.45 | 614 | 0.49 | 0.84 | 0.68 | 0.31 | 0.25 | 0.38 |
| Psychiatric conditions |  |  |  |  |  |  |  |  |  |  |
| Any anxiety disorder | 17,884 | 5.45 | 6.70 | 401 | 11.92 | 11.87 | 0.93 | 6.46 | 5.24 | 7.68 |
| Depression | 17,891 | 0.94 | 2.31 | 407 | 3.06 | 4.50 | 0.87 | 2.10 | 1.65 | 2.54 |
| Separation anxiety disorder | 17,622 | 1.55 | 2.27 | 390 | 3.33 | 3.61 | 0.77 | 1.79 | 1.40 | 2.17 |
| Panic disorder | 17,794 | 0.64 | 1.40 | 393 | 1.70 | 2.99 | 0.72 | 1.05 | 0.74 | 1.36 |
| Generalized anxiety disorder | 17,856 | 1.24 | 2.20 | 401 | 2.78 | 3.63 | 0.68 | 1.53 | 1.16 | 1.90 |
| Sleep problems | 30,179 | 0.11 | 0.34 | 616 | 0.35 | 0.60 | 0.67 | 0.23 | 0.18 | 0.28 |
| School phobia<sup>a</sup> | 17,855 | 0.23 | 0.74 | 396 | 0.73 | 1.38 | 0.65 | 0.50 | 0.36 | 0.63 |
| Social phobia | 17,869 | 1.77 | 2.61 | 410 | 3.40 | 3.68 | 0.62 | 1.63 | 1.26 | 2.00 |

Notes. Sex and birth year were included as covariates in the regression models (results not displayed). q-values were <p.0001 for all outcomes. <sup>a</sup> denotes outcomes that were only treated continuously, but not dichotomously.
ADHD, attention deficit hyperactivity disorder; ARFID, avoidant restrictive food intake disorder; A-TAC, Autism-Tics, ADHD, and other Comorbidities inventory; RRBI, restricted/repetitive behavior & interests; SCARED, Screen for Child Anxiety Related Emotional Disorders; SMFQ, Short Mood and Feelings Questionnaire. Std. B, standardized regression coefficient; Unstd. B, unstandardized regression coefficient; 95% CI LL/UL, lower/upper limits of the cluster-robust 95% confidence interval of the unstandardized regression coefficient.

**Table 4.** Logistic regression models for dichotomous outcomes on ARFID.

| Outcome | No ARFID |  |  | ARFID |  |  | OR | 95% CI<sub>LL</sub> | 95% CI<sub>UL</sub> |
| --- | --- | --- | --- | --- | --- | --- | --- | --- | --- |
|  | N | n | % | N | n | % |  |  |  |
| Neurodevelopmental conditions |  |  |  |  |  |  |  |  |  |
| Autism | 30,092 | 276 | 0.9 | 611 | 74 | 12.1 | 13.71 | 10.30 | 18.25 |
| ADHD | 30,068 | 582 | 1.9 | 608 | 103 | 16.9 | 9.43 | 7.37 | 12.07 |
| Perception problems | 30,179 | 1,043 | 3.5 | 616 | 136 | 22.1 | 7.55 | 6.12 | 9.31 |
| Learning disorders | 30,128 | 449 | 1.5 | 616 | 58 | 9.4 | 6.82 | 5.08 | 9.17 |
| Oppositional defiant disorder | 30,099 | 963 | 3.2 | 614 | 119 | 19.4 | 6.80 | 5.45 | 8.48 |
| DCD<sup>a</sup> | 30,128 | 505 | 1.7 | 616 | 64 | 10.4 | 6.66 | 5.02 | 8.83 |
| Conduct disorder | 30,105 | 319 | 1.1 | 613 | 36 | 5.9 | 5.28 | 3.60 | 7.75 |
| Tic disorders | 30,106 | 1,013 | 3.4 | 614 | 91 | 14.8 | 4.56 | 3.58 | 5.82 |
| Psychiatric conditions |  |  |  |  |  |  |  |  |  |
| OCD<sup>a</sup> | 30,126 | 593 | 2.0 | 616 | 68 | 11.0 | 5.87 | 4.45 | 7.73 |
| Generalized anxiety disorder | 17,856 | 474 | 2.7 | 401 | 51 | 12.7 | 5.24 | 3.78 | 7.28 |
| Depression | 17,891 | 544 | 3.0 | 407 | 53 | 13.0 | 4.57 | 3.36 | 6.22 |
| Panic disorder | 17,794 | 678 | 3.8 | 393 | 60 | 15.3 | 4.45 | 3.29 | 6.01 |
| Any anxiety disorder | 17,884 | 1,756 | 9.8 | 401 | 123 | 30.7 | 4.08 | 3.24 | 5.13 |
| Social phobia | 17,869 | 534 | 3.0 | 410 | 46 | 11.2 | 4.06 | 2.92 | 5.65 |
| Sleep problems | 30,179 | 1,679 | 5.6 | 616 | 123 | 20.0 | 4.04 | 3.26 | 5.00 |
| Separation anxiety disorder | 17,622 | 1,733 | 9.8 | 390 | 113 | 29.0 | 3.79 | 2.99 | 4.79 |
| Visual hallucinations<sup>a</sup> | 29,466 | 611 | 2.1 | 605 | 42 | 6.9 | 3.33 | 2.40 | 4.63 |

Notes. Sex and birth year were included as covariates in the regression models (results not displayed). q-values were <p.0001 for all outcomes. <sup>a</sup> denotes outcomes that were only treated dichotomously but not continuously.
ADHD, attention deficit hyperactivity disorder; ARFID, avoidant restrictive food intake disorder; A-TAC, Autism-Tics, ADHD, and other Comorbidities inventory; DCD, developmental coordination disorder; ODD, oppositional defiant disorder; OR, odds ratio; RRBI, restricted/repetitive behavior & interests; SCARED, Screen for Child Anxiety Related Emotional Disorders; SMFQ, Short Mood and Feelings Questionnaire. 95% CI LL/UL, lower/upper limits of the cluster-robust 95% confidence interval of the odds ratio.

ARFID was a highly significant predictor of higher mean symptom scores and higher odds of screening diagnosis for *all* measured outcomes. The highest effect size was seen for autism: children with ARFID had 14-fold odds of exceeding the cutoff for a screening diagnosis of autism compared with children without ARFID (odds ratio [OR]=13.71, 95% CI 10.30–18.25). This was followed by ADHD, for which children with ARFID had 9-fold odds (OR=9.43, 95% CI 7.37–12.07). Odds of other NDCs ranged from 7.55 (perception problems) to 4.56 (tic disorders). Odds of psychiatric conditions were 3- to 6-fold in the ARFID group, with the highest risk increase for OCD (OR=5.87, 95% CI 4.45–7.73) and GAD (OR=5.24, 95% CI 3.78–7.28).

The ARFID x sex interaction term was non-significant for all but one outcome (autism– restricted/repetitive behavior & interests; *q*=0.016; **Tables S1 & S2**). Sex had a highly significant main effect for most outcomes, with boys having more NDCs, depressive symptoms, and sleep problems, and girls having higher odds of anxiety disorders (**Tables S1 & S2**).

## Discussion

The current study is one of few epidemiological studies on ARFID comorbidity in a large general population sample comparing children with and without the ARFID phenotype. We examined symptom scores and odds of a broad range of co-occurring NDCs and psychiatric conditions assessed via parent report questionnaires. All analyzed symptom scores were significantly higher in children with ARFID than in children without ARFID. Furthermore, the ARFID group had significantly increased odds of a screening diagnosis for all analyzed conditions. A full 62% of children with ARFID had at least one screening diagnosis compared to 27% of children without ARFID.

### Neurodevelopmental conditions

Consistent with prior research, our findings revealed higher odds of NDCs among children with ARFID, with autism (OR∼14) and ADHD (OR∼10) exhibiting the highest increased odds. The prevalence rates of screening diagnoses of autism (12.1%) and ADHD (16.9%) among children with ARFID align with the lower to middle range of previously reported estimates (autism: 3–66%, ADHD: 3–23%), which is likely due to that we examined a population-based sample.^14,15,18,31,32^

Additionally, we identified increased odds of perception problems, learning disorders, OCD, tic disorders, conduct disorder, DCD, and sleep problems in the ARFID group. The increased prevalence of perception problems—here including sensitivity to touch and sounds and impaired proprioception—is likely explained by the fact that (1) sensory sensitivity to taste and smell is an integral part of ARFID and was used to define the ARFID phenotype in this sample and (2) sensory sensitivities among different senses are highly correlated.^33^ The prevalence of learning disorders in children with ARFID was found to be 9.4%, which is

consistent with prior research.^18,32^ Similarly, higher odds of OCD and sleep problems is supported by prior research.^34,35^ A register-based study on the same sample did not find a significantly increased risk of clinical diagnoses of OCD in the ARFID group, which might be explained by lower statistical power in using registered diagnoses compared to using parent reports.^11^ Additionally, in Sweden, OCD is often treated in primary care and that study did not include diagnoses received in primary care. Our findings of increased risks of tic disorder, conduct disorder, and DCD in children with ARFID, however, align well with what was seen in registered clinical diagnoses.^11^

Our analysis is the first to reveal a significantly higher odds of ODD in children with ARFID: 19.4% had a screening diagnosis of ODD, while previous research found a much lower prevalence of ODD in ARFID.^14^ Additionally, we observed a significantly higher risk of visual hallucinations in children with ARFID. To our knowledge, the only prior mention of this comorbidity was a case report of a patient with ARFID and Gitelman syndrome.^36^ **Anxiety and depression**

A screening diagnosis of anxiety disorder was present in 31% of children with ARFID (compared to 10% in children without ARFID), which aligns with previous findings reporting anxiety disorders in 35% to 72% of ARFID patients.^13–15,18,32,37^ Our observed prevalence falls toward the lower end of this range, which is again likely due to our non-clinical sample. Furthermore, previous studies have shown that anxiety symptoms are higher in patients with the fear profile than in patients with the sensory or low appetite profiles.^38^ The approach we used to identify ARFID preferentially captures the sensory-based profile, meaning that the fear of aversive consequences profile might be underrepresented in our sample.^5^ Consequently, our prevalence estimates potentially underestimate the association of ARFID and anxiety symptoms for the entire ARFID population.

In contrast to prior research, we did not find GAD to be the most prevalent anxiety presentation in ARFID; rather, separation anxiety was most common (29%). Earlier prevalence estimates for co-occurring GAD ranged from 18% to 50%, whereas separation anxiety was previously found to co-occur in 0–1% of patients with ARFID.^14,18^ The current finding is likely explained by that fact that separation anxiety is characterized by an earlier onset than GAD^39^ and that prior studies analyzed samples with an older mean age compared to ours.^14,18,31^ It is also possible that clinicians tend to diagnose anxiety symptoms of similar nature as separation anxiety in children and as GAD in adults. This could additionally have contributed to the lower prevalence of separation anxiety and the higher prevalence of GAD in prior studies, particularly those that relied on clinical diagnoses rather than questionnaire-based assessments. A hypothesis worth further investigation is that the increased prevalence of separation anxiety in ARFID may be explained by intertwined symptoms such that children fear separation from caregivers due to the possibility of unanticipated exposure to feared foods or situations related to their feeding disorder.

Moreover, the current study provides new evidence of increased symptoms of school phobia in children with ARFID. Increased school phobia may partly be attributed to concerns regarding mealtimes at school due to the restricted variety and/or quantity of food intake which may not be accommodated by the school and potentially elicit teasing by peers.^40^ Importantly, anxiety disorders may worsen ARFID symptoms by increasing avoidance behaviors (e.g., low willingness to try new foods) thereby acting as a maintaining mechanism of ARFID.^41^

In the current study, a screening diagnosis of depression in children with ARFID was less common than a screening diagnosis of anxiety disorder (13% vs. 31%), which is consistent with prior research showing the same pattern.^12,14,15^ Our findings also align with studies that have observed a lower prevalence of depression in ARFID compared to other eating disorders.^15^ The reason for this is not entirely clear, but may partly be related to age: on average, ARFID patients are younger than patients with other eating disorders,^12^ and mood disorders have a later average age of onset than anxiety disorders,^42^ leading to relatively lower prevalence of co-occurring depression in ARFID. Furthermore, in other eating disorders, weight and shape concerns exert a strong influence on negative self-evaluation^43^ which in turn is associated with depressive symptoms.^44^ The lack of body image concerns influencing self-evaluation in ARFID might contribute to the lower rate of depressive symptoms.

### Clinical implications

The current study provides insight into the neurodevelopmental and psychiatric profile of children with the ARFID phenotype. The results of this epidemiological study raise important issues related to the assessment and treatment of ARFID. Providers are recommended to introduce screening for NDCs, anxiety, and depression into ARFID assessment to facilitate early intervention and improve chances of successful recovery. A better understanding of how ARFID relates to other conditions can also facilitate tailoring treatment approaches that target not only ARFID but also co-occurring symptoms. Although various psychological treatments, including cognitive-behavioral therapy and family-based treatment, have been shown to reduce ARFID symptoms, these results are based on clinical case series and small-scale pilot randomized clinical trials.^45^ Psychological and pharmacological treatments that intentionally target ARFID and co-occurring symptoms simultaneously remain to be developed.^45^

### Strengths and limitations

The current study benefited from a large cohort of ∼31,000 children, including 616 children, who, by combining information from a variety of data sources met algorithmized diagnostic criteria for ARFID. Unlike most of prior research that investigated a narrow range of co-occurring conditions in relatively small clinical samples and often lacked control groups, our study examined a broad range of conditions in a non-clinical sample, reducing referral bias and therefore providing a more accurate reflection of the heterogeneous clinical presentations in a population of children with the ARFID phenotype, including those who are not actively seeking treatment or who lack access to it. By assessing co-occurring conditions both as symptom scores and as screening diagnoses, we offered a more comprehensive understanding of the health care needs in children with ARFID in a non-clinical child population. Given that ARFID is not included in ICD-10, and given the scarcity of treatment options for ARFID in Sweden, our results are indeed valuable for resource planning and foreshadow the complex needs that will have to be addressed in developing adequate treatments. Lastly, we used well-established measurement tools, aligned with DSM-5 criteria, enhancing the validity of our results.

The following limitations need to be considered. Despite our large sample size, generalizability of our findings is uncertain, since assessment of outcomes only relied on parent reports at child age 9 or 12, limiting extrapolation to adolescents and adults and potentially introducing rater bias and correlated measurement error.^46^ The current data are however complemented by our previous study in the same sample examining register-based clinical diagnoses from birth to age 18.^11^ Second, the response rate was 69% and it has been shown that responders in CATSS tended to be more healthy than non-responders.^22^ This could potentially have led to an underestimation of differences between the groups of children with and without ARFID. Furthermore, most children identified with the ARFID phenotype met criteria via low weight and psychosocial impairment, while only few were identified based on nutritional deficiencies and supplement dependence.^5^ Although this pattern has been observed in previous population-based screening studies,^6^ the rates of nutritional deficiencies and supplement dependence in the current sample were much lower than in studies examining clinical cohorts,^47^ indicating that our observed symptom pattern may be representative of ARFID in the general population, but not of ARFID in current clinical populations. In addition, our approach to identify the ARFID phenotype predominantly focused on the sensory-based avoidance component, often linked to selective eating.^48^ This may have resulted in a higher observed co-occurrence of NDCs and a lower co-occurrence of anxiety in the current sample. Future research could focus on longitudinal studies to examine the temporal relationship of ARFID with NDCs and psychiatric conditions. Gaining understanding of the order in which symptoms arise could provide valuable insight into the origins of ARFID. Additionally, future studies should examine common etiological (i.e., genetic and environmental) factors between ARFID and co-occurring conditions to shed light on *why* ARFID often coexists with NDCs and psychiatric conditions.

## Conclusion

The current study provides evidence for the frequent co-occurrence of a broad range of NDCs and psychiatric conditions with ARFID in the largest, non-clinical sample of children with and without the ARFID phenotype to date. Our results highlight the significant health care needs in children with ARFID due to multiple co-existing conditions. It is essential to implement screening of multiple potentially co-occurring symptoms in the assessment of ARFID to facilitate adequate referrals, reduce duration of illness exposure, and contribute to successful recovery.

## Disclosures

### Authors’ contributions

Conceptualization: Lisa Dinkler, Manda Nyholmer Data curation: Lisa Dinkler, Manda Nyholmer

Formal analysis: Lisa Dinkler, Manda Nyholmer, Marie-Louis Wronski Funding acquisition: Lisa Dinkler, Manda Nyholmer, Cynthia M. Bulik

Investigation: Manda Nyholmer, Lisa Dinkler, Paul Lichtenstein, Sebastian Lundström, Henrik Larsson

Methodology: Lisa Dinkler, Manda Nyholmer, Ralf Kuja-Halkola, Mark J. Taylor Project administration: Lisa Dinkler

Resources: Paul Lichtenstein, Sebastian Lundström, Henrik Larsson

Software: Lisa Dinkler, Manda Nyholmer, Marie-Louis Wronski, Ralf Kuja-Halkola Supervision: Lisa Dinkler

Validation: Ralf Kuja-Halkola, Lisa Dinkler Visualization: Lisa Dinkler

Writing – original draft: Manda Nyholmer, Lisa Dinkler, Liv Hog Writing – review & editing: All authors

### Data sharing

Swedish register data and data from the Child and Adolescent Twin study in Sweden are not publicly available and cannot be shared.

