## Supplemental file for "Neurodevelopmental and psychiatric conditions in 600 Swedish Children with the ARFID phenotype"

| **Table S1**. Linear regression models for continuous outcomes on ARFID, including the ARFID x sex interaction term | | | | | | | | | | | | |
| --- | --- | --- | --- | --- | --- | --- | --- | --- | --- | --- | --- | --- |
| **Outcome** | **Term** | **Unstd. B** | | | | **95% CI_LL_** | **95% CI_UL_** | **Robust SE** | ***t statistic*** | | ***p-value*** | ***q-value*** |
| *Neurodevelopmental conditions* | | | |  | | |  |  | |  |  |  |
| Autism - Total | | |  |  | | |  |  | |  |  |  |
|  | ARFID | | 2.75 | 2.34 | | | 3.16 | 0.21 | | 13.15 | <0.0001 | <0.0001 |
|  | Female | | -0.35 | -0.39 | | | -0.32 | 0.02 | | -18.59 | <0.0001 | <0.0001 |
|  | ARFID x female | | -0.80 | -1.40 | | | -0.20 | 0.31 | | -2.61 | 0.009 | 0.085 |
| Autism - Social communication^a^ | | |  | | | |  |  | |  |  |  |
|  | ARFID | | 1.70 | | 1.42 | | 1.98 | 0.14 | | 12.00 | <0.0001 | <0.0001 |
|  | Female | | -0.21 | | -0.23 | | -0.18 | 0.01 | | -15.38 | <0.0001 | <0.0001 |
|  | ARFID x female | | -0.42 | | -0.83 | | -0.01 | 0.21 | | -1.98 | 0.047 | 0.162 |
| Autism - RRBI^a^ | | |  | | | |  |  | |  |  |  |
|  | ARFID | | 1.05 | | 0.89 | | 1.21 | 0.08 | | 12.98 | <0.0001 | <0.0001 |
|  | Female | | -0.15 | | -0.16 | | -0.13 | 0.01 | | -19.56 | <0.0001 | <0.0001 |
|  | ARFID x female | | -0.38 | | -0.61 | | -0.16 | 0.11 | | -3.33 | 0.0009 | 0.016 |
| ADHD - Total | | |  | | | |  |  | |  |  |  |
|  | ARFID | | 4.02 | | 3.45 | | 4.59 | 0.29 | | 13.82 | <0.0001 | <0.0001 |
|  | Female | | -0.89 | | -0.97 | | -0.82 | 0.04 | | -23.82 | <0.0001 | <0.0001 |
|  | ARFID x female | | -0.89 | | -1.78 | | 0.00 | 0.45 | | -1.95 | 0.051 | 0.162 |
| ADHD - Inattention^a^ | | |  | | | |  |  | |  |  |  |
|  | ARFID | | 2.14 | | 1.83 | | 2.45 | 0.16 | | 13.61 | <0.0001 | <0.0001 |
|  | Female | | -0.50 | | -0.54 | | -0.46 | 0.02 | | -23.81 | <0.0001 | <0.0001 |
|  | ARFID x female | | -0.34 | | -0.83 | | 0.15 | 0.25 | | -1.37 | 0.170 | 0.429 |
| ADHD - Impulsivity/hyperactivity^a^ | | |  | | | |  |  | |  |  |  |
|  | ARFID | | 1.87 | | 1.56 | | 2.18 | 0.16 | | 11.87 | <0.0001 | <0.0001 |
|  | Female | | -0.39 | | -0.43 | | -0.35 | 0.02 | | -19.24 | <0.0001 | <0.0001 |
|  | ARFID x female | | -0.54 | | -1.00 | | -0.07 | 0.24 | | -2.24 | 0.025 | 0.118 |
| Learning disorders | | |  | | | |  |  | |  |  |  |
|  | ARFID | | 0.55 | | 0.43 | | 0.66 | 0.06 | | 9.57 | <0.0001 | <0.0001 |
|  | Female | | -0.05 | | -0.07 | | -0.04 | 0.01 | | -7.08 | <0.0001 | <0.0001 |
|  | ARFID x female | | -0.08 | | -0.25 | | 0.10 | 0.09 | | -0.88 | 0.378 | 0.553 |
| Tic disorders | | |  | | | |  |  | |  |  |  |
|  | ARFID | | 0.38 | | 0.28 | | 0.47 | 0.05 | | 7.69 | <0.0001 | <0.0001 |
|  | Female | | -0.10 | | -0.11 | | -0.09 | 0.01 | | -18.61 | <0.0001 | <0.0001 |
|  | ARFID x female | | -0.16 | | -0.29 | | -0.03 | 0.07 | | -2.43 | 0.015 | 0.096 |
| ODD | | |  | | | |  |  | |  |  |  |
|  | ARFID | | 0.90 | | 0.75 | | 1.06 | 0.08 | | 11.54 | <0.0001 | <0.0001 |
|  | Female | | -0.13 | | -0.15 | | -0.11 | 0.01 | | -12.46 | <0.0001 | <0.0001 |
|  | ARFID x female | | -0.09 | | -0.32 | | 0.15 | 0.12 | | -0.73 | 0.466 | 0.632 |
| Conduct disorder | | |  | | | |  |  | |  |  |  |
|  | ARFID | | 0.28 | | 0.19 | | 0.37 | 0.05 | | 6.10 | <0.0001 | <0.0001 |
|  | Female | | -0.04 | | -0.05 | | -0.04 | 0.00 | | -9.90 | <0.0001 | <0.0001 |
|  | ARFID x female | | -0.08 | | -0.20 | | 0.05 | 0.06 | | -1.21 | 0.226 | 0.429 |
| Perception problems | | |  | | | |  |  | |  |  |  |
|  | ARFID | | 0.59 | | 0.49 | | 0.70 | 0.05 | | 11.06 | <0.0001 | <0.0001 |
|  | Female | | -0.04 | | -0.06 | | -0.03 | 0.01 | | -7.97 | <0.0001 | <0.0001 |
|  | ARFID x female | | -0.08 | | -0.25 | | 0.08 | 0.08 | | -0.99 | 0.323 | 0.523 |
| **Table S1** (continued) | | |  | |  | |  |  | |  |  |  |
| **Outcome** | **Term** | | **Unstd. B** | **95% CI_LL_** | | | **95% CI_UL_** | **Robust SE** | | ***t statistic*** | ***p-value*** | ***q-value*** |
| *Psychiatric conditions* | | |  | |  | |  |  | |  |  |  |
| Depression | | |  | | | |  |  | |  |  |  |
|  | ARFID | | 2.22 | | 1.61 | | 2.83 | 0.31 | | 7.19 | <0.0001 | <0.0001 |
|  | Female | | -0.15 | | -0.22 | | -0.08 | 0.04 | | -4.06 | <0.0001 | <0.0001 |
|  | ARFID x female | | -0.29 | | -1.18 | | 0.60 | 0.45 | | -0.64 | 0.520 | 0.658 |
| Any anxiety disorder | | |  | | | |  |  | |  |  |  |
|  | ARFID | | 6.71 | | 4.99 | | 8.42 | 0.87 | | 7.68 | <0.0001 | <0.0001 |
|  | Female | | 0.66 | | 0.45 | | 0.87 | 0.11 | | 6.08 | <0.0001 | <0.0001 |
|  | ARFID x female | | -0.59 | | -3.04 | | 1.87 | 1.25 | | -0.47 | 0.640 | 0.760 |
| Panic disorder | | |  | | | |  |  | |  |  |  |
|  | ARFID | | 1.04 | | 0.66 | | 1.43 | 0.20 | | 5.29 | <0.0001 | <0.0001 |
|  | Female | | 0.00 | | -0.04 | | 0.04 | 0.02 | | -0.06 | 0.954 | 0.954 |
|  | ARFID x female | | 0.03 | | -0.62 | | 0.67 | 0.33 | | 0.08 | 0.938 | 0.939 |
| Generalized anxiety disorder | | |  | | | |  |  | |  |  |  |
|  | ARFID | | 1.54 | | 1.04 | | 2.04 | 0.25 | | 6.07 | <0.0001 | <0.0001 |
|  | Female | | 0.16 | | 0.09 | | 0.23 | 0.04 | | 4.62 | <0.0001 | <0.0001 |
|  | ARFID x female | | -0.03 | | -0.78 | | 0.72 | 0.38 | | -0.08 | 0.939 | 0.939 |
| Separation anxiety disorder | | |  | | | |  |  | |  |  |  |
|  | ARFID | | 1.99 | | 1.44 | | 2.54 | 0.28 | | 7.09 | <0.0001 | <0.0001 |
|  | Female | | 0.21 | | 0.14 | | 0.29 | 0.04 | | 5.80 | <0.0001 | <0.0001 |
|  | ARFID x female | | -0.48 | | -1.23 | | 0.27 | 0.38 | | -1.25 | 0.212 | 0.429 |
| Social phobia^a^ | | |  | | | |  |  | |  |  |  |
|  | ARFID | | 1.59 | | 1.08 | | 2.11 | 0.26 | | 6.04 | <0.0001 | <0.0001 |
|  | Female | | 0.26 | | 0.17 | | 0.34 | 0.04 | | 6.06 | <0.0001 | <0.0001 |
|  | ARFID x female | | 0.09 | | -0.64 | | 0.82 | 0.37 | | 0.24 | 0.813 | 0.908 |
| School phobia | | |  | | | |  |  | |  |  |  |
|  | ARFID | | 0.57 | | 0.37 | | 0.77 | 0.10 | | 5.68 | <0.0001 | <0.0001 |
|  | Female | | 0.03 | | 0.01 | | 0.05 | 0.01 | | 2.58 | 0.01 | 0.011 |
|  | ARFID x female | | -0.18 | | -0.45 | | 0.10 | 0.14 | | -1.26 | 0.206 | 0.429 |
| Sleep problems | | |  | | | |  |  | |  |  |  |
|  | ARFID | | 0.25 | | 0.19 | | 0.32 | 0.03 | | 7.55 | <0.0001 | <0.0001 |
|  | Female | | -0.02 | | -0.03 | | -0.01 | 0.00 | | -4.47 | <0.0001 | <0.0001 |
|  | ARFID x female | | -0.05 | | -0.14 | | 0.05 | 0.05 | | -0.97 | 0.330 | 0.523 |
| *Notes.* Birth year was included as a covariate in the regression models (results not displayed). ^a^ denotes outcomes that were only treated dichotomously but not continuously. Highlighting: statistically significant interaction term (pink), statistically significantly higher symptoms scores in females (green), statistically significantly lower symptoms scores in females (blue). ADHD, attention deficit hyperactivity disorder; ARFID, avoidant restrictive food intake disorder; A-TAC, Autism-Tics, ADHD, and other Comorbidities inventory; Robust SE, cluster-robust standard error; RRBI, restricted/repetitive behavior & interests; SCARED, Screen for Child Anxiety Related Emotional Disorders; SMFQ, Short Mood and Feelings Questionnaire. Unstd. B, unstandardized regression coefficient; 95% CI LL/UL, lower/upper limits of the cluster-robust 95% confidence interval of the unstandardized regression coefficient. | | | | | | | | | | | | |

| **Table S2**. Logistic regression models for dichotomous outcomes on ARFID, including the ARFID x sex interaction term | | | | | | | | | | | | | | | |
| --- | --- | --- | --- | --- | --- | --- | --- | --- | --- | --- | --- | --- | --- | --- | --- |
| **Outcome** | **Term** | | **OR** | | **95% CI_LL_** | | **95% CI_UL_** | | **Robust SE** | | ***Z* statistic** | | ***p*-value** | | ***q*-value** |
| *Neurodevelopmental conditions* | |  | |  | |  | |  | |  | |  | |  | |
| Autism | |  | |  | |  | |  | |  | |  | |  | |
|  | ARFID | | 12.69 | | 9.12 | | 17.67 | | 0.17 | | 15.07 | | <0.0001 | | <0.0001 |
|  | Female | | 0.40 | | 0.31 | | 0.53 | | 0.14 | | -6.55 | | <0.0001 | | <0.0001 |
|  | ARFID x female | | 1.33 | | 0.70 | | 2.52 | | 0.33 | | 0.87 | | 0.383 | | 0.810 |
| ADHD | |  | |  | |  | |  | |  | |  | |  | |
|  | ARFID | | 8.29 | | 6.20 | | 11.08 | | 0.15 | | 14.28 | | <0.0001 | | <0.0001 |
|  | Female | | 0.40 | | 0.33 | | 0.49 | | 0.10 | | -9.30 | | <0.0001 | | <0.0001 |
|  | ARFID x female | | 1.56 | | 0.93 | | 2.60 | | 0.26 | | 1.69 | | 0.091 | | 0.545 |
| Learning disorders | |  | |  | |  | |  | |  | |  | |  | |
|  | ARFID | | 6.96 | | 4.83 | | 10.02 | | 0.19 | | 10.42 | | <0.0001 | | <0.0001 |
|  | Female | | 0.93 | | 0.76 | | 1.13 | | 0.10 | | -0.77 | | 0.442 | | 0.468 |
|  | ARFID x female | | 0.95 | | 0.52 | | 1.73 | | 0.30 | | -0.17 | | 0.868 | | 0.952 |
| Tic disorders | |  | |  | |  | |  | |  | |  | |  | |
|  | ARFID | | 4.60 | | 3.51 | | 6.05 | | 0.14 | | 10.97 | | <0.0001 | | <0.0001 |
|  | Female | | 0.38 | | 0.33 | | 0.44 | | 0.07 | | -13.19 | | <0.0001 | | <0.0001 |
|  | ARFID x female | | 0.96 | | 0.53 | | 1.74 | | 0.30 | | -0.13 | | 0.899 | | 0.952 |
| ODD | |  | |  | |  | |  | |  | |  | |  | |
|  | ARFID | | 6.15 | | 4.67 | | 8.09 | | 0.14 | | 12.96 | | <0.0001 | | <0.0001 |
|  | Female | | 0.61 | | 0.53 | | 0.70 | | 0.07 | | -7.01 | | <0.0001 | | <0.0001 |
|  | ARFID x female | | 1.33 | | 0.85 | | 2.08 | | 0.23 | | 1.27 | | 0.205 | | 0.738 |
| Conduct disorder | |  | |  | |  | |  | |  | |  | |  | |
|  | ARFID | | 4.19 | | 2.53 | | 6.94 | | 0.26 | | 5.56 | | <0.0001 | | <0.0001 |
|  | Female | | 0.49 | | 0.38 | | 0.63 | | 0.13 | | -5.51 | | <0.0001 | | <0.0001 |
|  | ARFID x female | | 2.03 | | 0.95 | | 4.34 | | 0.39 | | 1.82 | | 0.069 | | 0.545 |
| Perception problems | |  | |  | |  | |  | |  | |  | |  | |
|  | ARFID | | 7.39 | | 5.72 | | 9.55 | | 0.13 | | 15.32 | | <0.0001 | | <0.0001 |
|  | Female | | 0.68 | | 0.59 | | 0.77 | | 0.07 | | -5.76 | | <0.0001 | | <0.0001 |
|  | ARFID x female | | 1.06 | | 0.68 | | 1.65 | | 0.22 | | 0.27 | | 0.790 | | 0.950 |
| DCD | |  | |  | |  | |  | |  | |  | |  | |
|  | ARFID | | 6.27 | | 4.45 | | 8.83 | | 0.17 | | 10.52 | | <0.0001 | | <0.0001 |
|  | Female | | 0.60 | | 0.50 | | 0.72 | | 0.09 | | -5.38 | | <0.0001 | | <0.0001 |
|  | ARFID x female | | 1.20 | | 0.66 | | 2.19 | | 0.31 | | 0.60 | | 0.547 | | 0.810 |
| *Psychiatric conditions* | |  | |  | |  | |  | |  | |  | |  | |
| Depression | |  | |  | |  | |  | |  | |  | |  | |
|  | ARFID | | 4.30 | | 2.95 | | 6.28 | | 0.19 | | 7.56 | | <0.0001 | | <0.0001 |
|  | Female | | 0.59 | | 0.49 | | 0.71 | | 0.09 | | -5.68 | | <0.0001 | | <0.0001 |
|  | ARFID x female | | 1.20 | | 0.63 | | 2.30 | | 0.33 | | 0.55 | | 0.585 | | 0.810 |
| Any anxiety disorder | |  | |  | |  | |  | |  | |  | |  | |
|  | ARFID | | 4.33 | | 3.19 | | 5.87 | | 0.16 | | 9.42 | | <0.0001 | | <0.0001 |
|  | Female | | 1.15 | | 1.04 | | 1.28 | | 0.05 | | 2.62 | | 0.009 | | 0.014 |
|  | ARFID x female | | 0.87 | | 0.55 | | 1.38 | | 0.24 | | -0.59 | | 0.552 | | 0.810 |
| Panic disorder | |  | |  | |  | |  | |  | |  | |  | |
|  | ARFID | | 4.61 | | 3.12 | | 6.80 | | 0.20 | | 7.69 | | <0.0001 | | <0.0001 |
|  | Female | | 0.98 | | 0.84 | | 1.15 | | 0.08 | | -0.21 | | 0.834 | | 0.834 |
|  | ARFID x female | | 0.92 | | 0.49 | | 1.72 | | 0.32 | | -0.26 | | 0.792 | | 0.950 |
| Generalized anxiety disorder | |  | |  | |  | |  | |  | |  | |  | |
|  | ARFID | | 5.74 | | 3.72 | | 8.84 | | 0.22 | | 7.92 | | <0.0001 | | <0.0001 |
|  | Female | | 1.12 | | 0.93 | | 1.36 | | 0.10 | | 1.19 | | 0.235 | | 0.264 |
|  | ARFID x female | | 0.81 | | 0.41 | | 1.58 | | 0.34 | | -0.62 | | 0.534 | | 0.810 |
| **Table S2.** (continued) | | |  | |  | |  | |  | |  | |  | |  |
| **Outcome** | **Term** | | **OR** | | **95% CI_LL_** | | **95% CI_UL_** | | **Robust SE** | | ***Z* statistic** | | ***p*-value** | | ***q*-value** |
| Separation anxiety disorder | |  | |  | |  | |  | |  | |  | |  | |
|  | ARFID | | 4.33 | | 3.20 | | 5.87 | | 0.16 | | 9.44 | | <0.0001 | | <0.0001 |
|  | Female | | 1.25 | | 1.12 | | 1.39 | | 0.05 | | 4.07 | | <0.0001 | | <0.0001 |
|  | ARFID x female | | 0.73 | | 0.45 | | 1.17 | | 0.24 | | -1.31 | | 0.19 | | 0.738 |
| Social phobia^a^ | |  | |  | |  | |  | |  | |  | |  | |
|  | ARFID | | 4.67 | | 3.03 | | 7.20 | | 0.22 | | 6.99 | | <0.0001 | | <0.0001 |
|  | Female | | 1.17 | | 0.98 | | 1.40 | | 0.09 | | 1.70 | | 0.088 | | 0.106 |
|  | ARFID x female | | 0.71 | | 0.36 | | 1.40 | | 0.35 | | -0.98 | | 0.327 | | 0.810 |
| School phobia | |  | |  | |  | |  | |  | |  | |  | |
|  | ARFID | | 7.64 | | 4.94 | | 11.80 | | 0.22 | | 9.15 | | <0.0001 | | <0.0001 |
|  | Female | | 1.25 | | 1.01 | | 1.54 | | 0.11 | | 2.03 | | 0.042 | | 0.064 |
|  | ARFID x female | | 0.45 | | 0.21 | | 0.93 | | 0.38 | | -2.14 | | 0.032 | | 0.545 |
| OCD | |  | |  | |  | |  | |  | |  | |  | |
|  | ARFID | | 6.30 | | 4.48 | | 8.85 | | 0.17 | | 10.60 | | <0.0001 | | <0.0001 |
|  | Female | | 0.86 | | 0.73 | | 1.02 | | 0.09 | | -1.74 | | 0.083 | | 0.106 |
|  | ARFID x female | | 0.81 | | 0.45 | | 1.47 | | 0.30 | | -0.68 | | 0.497 | | 0.810 |
| Visual hallucinations | |  | |  | |  | |  | |  | |  | |  | |
|  | ARFID | | 2.84 | | 1.84 | | 4.38 | | 0.22 | | 4.72 | | <0.0001 | | <0.0001 |
|  | Female | | 0.84 | | 0.71 | | 1.00 | | 0.09 | | -2.00 | | 0.046 | | 0.064 |
|  | ARFID x female | | 1.47 | | 0.76 | | 2.87 | | 0.34 | | 1.14 | | 0.254 | | 0.763 |
| Sleep problems | |  | |  | |  | |  | |  | |  | |  | |
|  | ARFID | | 4.02 | | 3.07 | | 5.26 | | 0.14 | | 10.14 | | <0.0001 | | <0.0001 |
|  | Female | | 0.81 | | 0.73 | | 0.90 | | 0.05 | | -4.09 | | <0.0001 | | <0.0001 |
|  | ARFID x female | | 1.01 | | 0.66 | | 1.56 | | 0.22 | | 0.05 | | 0.957 | | 0.957 |
| *Notes.* Birth year was included as a covariate in the regression models (results not displayed). ^a^ denotes outcomes that were only treated dichotomously but not continuously. Highlighting: statistically significantly higher risk in females (green), statistically significantly lower risk in females (blue). ADHD, attention deficit hyperactivity disorder; ARFID, avoidant restrictive food intake disorder; A-TAC, Autism-Tics, ADHD, and other Comorbidities inventory; DCD, developmental coordination disorder; ODD, oppositional defiant disorder; OR, odds ratio; Robust SE, cluster-robust standard error; RRBI, restricted/repetitive behavior & interests; SCARED, Screen for Child Anxiety Related Emotional Disorders; SMFQ, Short Mood and Feelings Questionnaire. 95% CI LL/UL, lower/upper limits of the cluster-robust 95% confidence interval of the odds ratio. | | | | | | | | | | | | | | | |
